# Improved Hemodynamic Performance and Reduced Paravalvular Regurgitation with the SAPIEN 3 Ultra RESILIA Valve: A Propensity-Matched Single-Center TAVR Study

**DOI:** 10.64898/2026.01.29.26345172

**Authors:** Meha Krishnareddigari, Suhas Yarra, Golsa Joodi, Radoslav I. Zinoviev, Olcay Aksoy

## Abstract

**Background:** Transcatheter aortic valve replacement (TAVR) has rapidly evolved into a standardized treatment for severe aortic stenosis, particularly in patients at increased surgical risk. The fifth-generation SAPIEN 3 Ultra RESILIA (S3UR) valve notably incorporates RESILIA-treated tissue as well as an enhanced external skirt in order to reduce structural valve deterioration (SVD) and paravalvular leak (PVL). However, real-world data on its clinical performance remains limited.

**Objectives:** To evaluate procedural, hemodynamic, and short-term clinical outcomes of the S3UR valve compared to earlier-generation SAPIEN 3 (S3) and SAPIEN 3 Ultra (S3U) platforms in Ronald Raegan UCLA medical center.

**Methods:** 513 patients who underwent transfemoral TAVR at Ronald Reagan UCLA Medical Center between 2022 and 2024 were analyzed. Of these, 216 received the S3UR valve and 297 received S3U/S3 valves. Propensity-score matching (1:1) yielded 181 well-balanced patient pairs. Primary endpoints included device success per VARC-3 criteria, with secondary endpoints encompassing 30-day safety, echocardiographic performance, and procedural complications.

**Results:** The S3UR group demonstrated significantly lower post-procedural and 30-day mean aortic valve gradients (7.45 ± 3.37 mmHg and 9.06 ± 2.94 mmHg, respectively; p < 0.001) compared to the S3U/S3 group. Rates of moderate or greater PVL were 0% in the S3UR group versus 8.9–10.1% in S3U/S3 patients (p < 0.001). Procedural success exceeded 98% in both groups, with no significant differences in stroke, mortality, or new pacemaker implantation. Readmission rates trended lower in the S3UR cohort (7.8% vs. 13.9%), though not statistically significant.

**Conclusions:** The SAPIEN 3 Ultra RESILIA valve demonstrated superior hemodynamic performance and significantly reduced PVL compared to earlier-generation balloon-expandable valves, while maintaining comparable safety and procedural success. These findings support the S3UR as a preferred TAVR platform in contemporary clinical practice, with ongoing follow-up needed to evaluate long-term durability.

## Introduction

Transcatheter aortic valve replacement (TAVR) has revolutionized the treatment of severe aortic stenosis, particularly in patients at intermediate or high surgical risk^1^. As we know it today, technological advances in transcatheter heart valve (THV) design have aimed to enhance procedural safety, improve hemodynamic performance, as well as extend long-term durability to mitigate human rick^2^. Among these developments the SAPIEN 3 Ultra Resilia (S3UR) represents the fifth-generation balloon-expandable THV platform in that it incorporates RESILIA-treated bovine pericardial leaflets; these are intended to reduce structural valve deterioration (SVD) and provide an enhanced external skirt to minimize paravalvular leak (PVL), particularly in larger valve sizes^3^.

The SAPIEN 3 Ultra Resilia (S3UR) valve introduces several key design enhancements over its predecessors, the SAPIEN 3 (S3) and SAPIEN 3 Ultra (S3U). Perhaps most notably, the S3UR incorporates RESILIA tissue technology, which essentially involves a proprietary preservation process that almost eliminates free aldehydes, compounds known to promote calcification, and then stabilizes the tissue to resist structural valve deterioration over the course of time^4^. This novel advancement is intended to increase long-term durability and reduce the likelihood of valve failure, often a critical consideration as TAVR expands to younger and lower-risk patient populations globally^5^. It is also important to note the S3UR 29-mm valve includes a taller and more conformable external skirt compared to the S3U and S3 platforms; this change is designed to improve annular sealing and reduce the incidence of paravalvular leak^6^. Furthermore, it should be considered that smaller valve sizes (20 mm and 23 mm) feature a redesigned commissural leaflet suspension that enhances leaflet mobility and hemodynamic performance by promoting more complete valve opening during systole^7^. These modifications, summarized in Figure 1 as well, represent a significant evolution in THV technology and thus aim to address limitations observed with earlier-generation valves.

**Figure 1.**
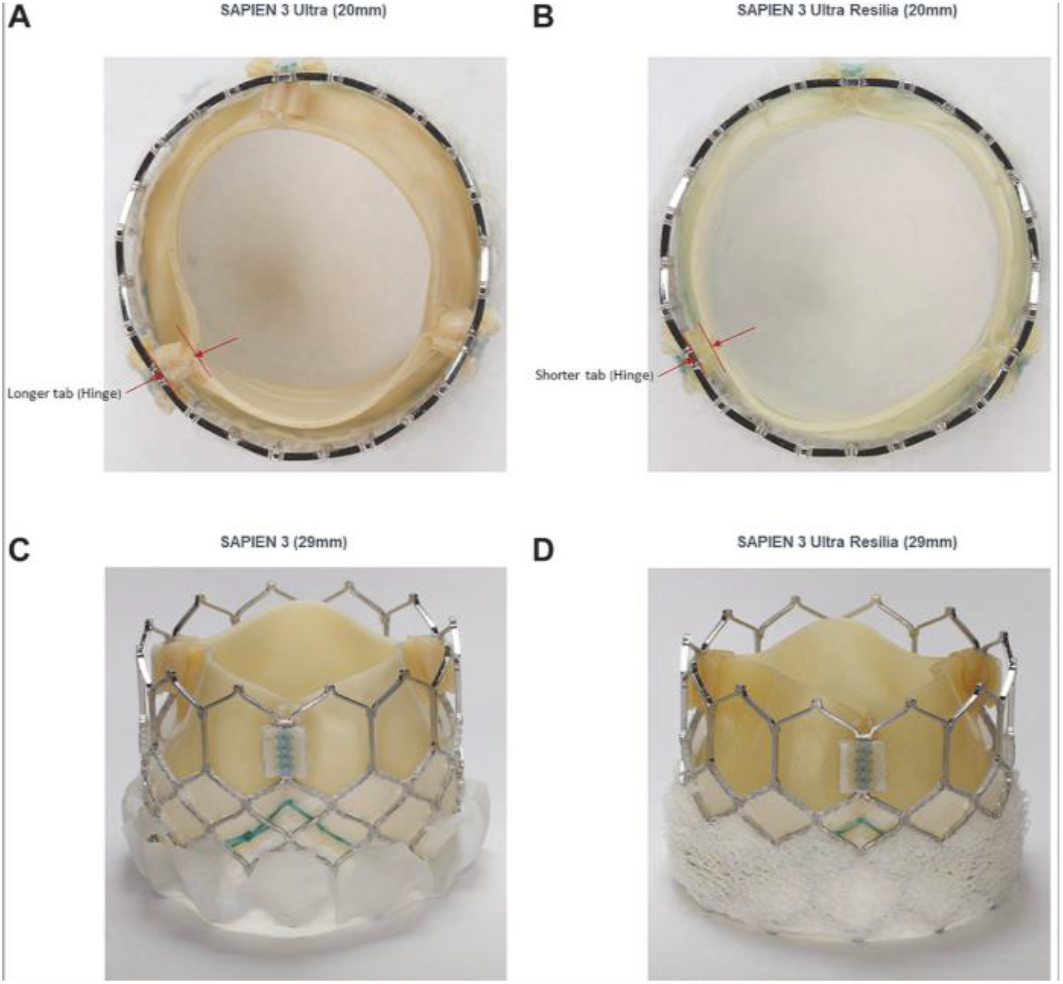
Illustrated are the design and structural elements of the 20-mm SAPIEN 3 Ultra (S3U) valve (A), 20-mm SAPIEN 3 Ultra Resilia (S3UR) valve (B), 29-mm SAPIEN 3 (S3) valve (C), and 29-mm S3UR valve (D). As depicted, the S3UR 20-mm valve features shorter commissural tab lengths compared to the S3U, potentially enhancing hemodynamic function within the body. Additionally, it is to note the S3UR 29-mm valve incorporates specialized leaflet tissue that is aimed at minimizing structural valve deterioration from calcification. Said leaflet also features an extended external skirt to better seal the annulus as well as reduce paravalvular leak.

### Structural Features of the S3UR, S3U, and S3 Transcatheter Heart Valves

Although multicenter data from national registries have demonstrated improved hemodynamics and similar safety outcomes with S3UR compared to S3 and S3U^8^, to date we know limited information on its real-world performance in single-center populations. In this study, we evaluate procedural, in-hospital, and 30-day outcomes among patients who underwent TAVR with the S3UR versus earlier-generation valve platforms at UCLA Ronald Reagan Medical Center between 2022 and early 202. Using propensity-score matching and statistical analysis to balance baseline characteristics, we aim to assess the safety, efficacy, and clinical impact of the S3UR valve in a contemporary, real-world setting such as a high-performance medical center. This study therefore creates an opportunity to evaluate and compare clinical outcomes, echocardiographic performance, as well as patient characteristics between S3UR and prior-generation balloon-expandable valves such as the SAPIEN 3 (S3) and SAPIEN 3 Ultra (S3U) in a real-world, single-center cohort.

## Methods

### Study Population

This single-center and retrospective study was approved by the UCLA Institutional Review Board with a waiver of informed consent due to the minimal risk design. All patients who underwent transfemoral TAVR using either the Sapien 3 Ultra Resilia (S3UR) or Sapien 3 Ultra/Sapien 3 (S3U/S3) valve platforms at Ronald Reagan UCLA Medical Center between 2022 and 2024 were included. Patients were excluded if they had a prior surgical or transcatheter aortic valve prosthesis, underwent valve-in-valve procedures, or had non-transfemoral access. A total of 513 patients met the inclusion criteria, with 216 patients receiving the S3UR valve and 297 receiving the S3U/S3 valve. As depicted in figure 2, propensity-score matching was used to generate two well-balanced cohorts of 181 patients each for comparative analysis of clinical and echocardiographic outcomes.

**Figure 2.**
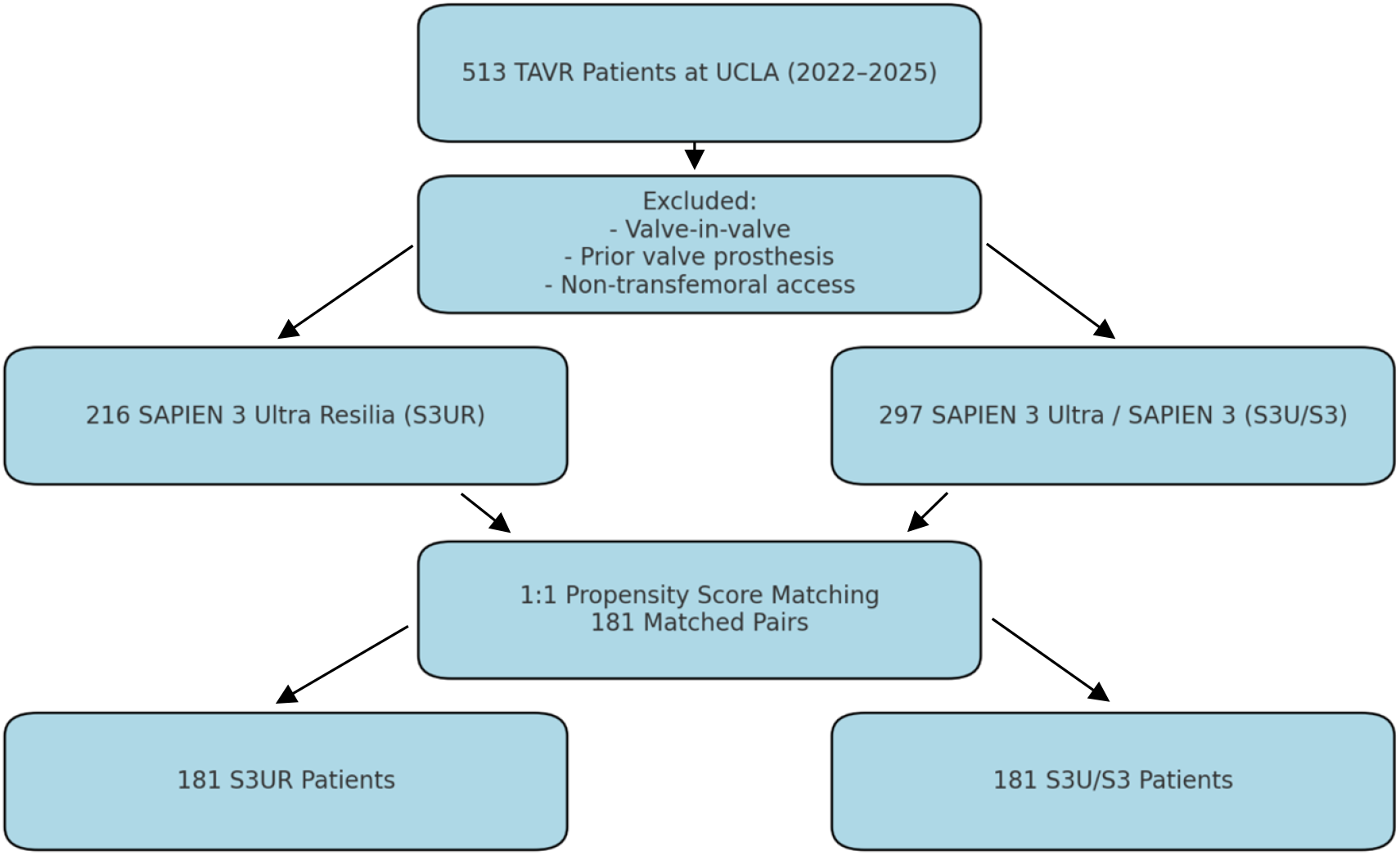
Study flow diagram showing derivation of matched cohorts for single-center analysis at UCLA. Patients undergoing transfemoral TAVR between 2022 and 2024 were screened. Exclusion criteria included prior valve prosthesis, valve-in-valve procedures, and non-transfemoral access. Propensity score matching resulted in 181 patient pairs treated with either the SAPIEN 3 Ultra RESILIA (S3UR) or SAPIEN 3 / SAPIEN 3 Ultra (S3U/S3) valve platforms.

### Valve Design and Delivery System

All transcatheter heart valves utilized at Ronald Raegan were manufactured by Edwards Lifesciences. The S3UR valve maintains the frame design of the S3U/S3 platform but incorporates some advanced tissue preservation technology to reduce calcification including the elimination of free aldehydes and the use of a proprietary leaflet treatment. In addition, the 20- and 23-mm S3UR valves also feature a revised commissural leaflet suspension designed to optimize leaflet kinematics. All S3U, S3, and S3UR valves were surgically implanted using the Commander Delivery System and eSheath introducer to minimize patient risk

### Endpoints

The primary endpoint as defined was device success as defined by the Valve Academic Research Consortium-3 (VARC-3) criteria; secondary endpoints included early safety at 30 days, procedural complications, and in-hospital echocardiographic outcomes as depicted in figures to follow. It is to note hemodynamic measurements were derived from site-reported transthoracic echocardiograms performed on each patient undergoing TAVR prior to discharge. Lastly, clinical endpoints as well as data definitions followed institutional standards aligned with STS/ACC TVT Registry guidelines.

### Propensity-Score Matching and Statistical Analysis

To best mitigate baseline differences between treatment groups, patients treated with the S3UR valve were propensity-score matched 1:1 to the patients that were treated with S3U/S3 using a nearest-neighbor algorithm without alternative replacement. Statistical propensity matching was performed utilizing 21 baseline clinical and echocardiographic covariates, with some examples involving demographics, comorbidities, STS risk score, valve characteristics, and functional status. Additionally, missing baseline data were imputed using Markov Chain Monte Carlo multiple imputation within the same dataset. To ensure accuracy of data, a caliper width of 0.02 times the standard deviation of the logit of the propensity score was applied and balance between groups was assessed using absolute standardized differences (ASDs). Values <0.1 were in this case considered indicative of good balance.

Next, continuous variables were summarized using mean ± standard deviation or median with interquartile ranges, and then compared using two-sample t-tests or Wilcoxon rank-sum tests as appropriate in each given scenario. Thus, categorical variables were summarized as counts and percentages and compared utilizing chi-square or Fisher’s exact tests. In terms of further data analysis, Kaplan-Meier estimates were used to calculate 30-day event rates with comparisons made utilizing the log-rank test. In this case, a two-sided p-value <0.05 was considered statistically significant. All analyses were performed using SAS version 9.4 (SAS Institute, Cary, NC).

## Results

### Baseline Characteristics

A total of 513 patients at Ronald Raegan underwent transfemoral transcatheter aortic valve replacement (TAVR) between 2022 and 2024. Of these we know 216 patients received the SAPIEN 3 Ultra Resilia (S3UR) valve, while 297 patients were treated with either the SAPIEN 3 Ultra (S3U) or SAPIEN 3 (S3) valves. In order to enable a more rigorous comparison of clinical outcomes across platforms statistical propensity-score matching was performed using 37 baseline clinical and echocardiographic variables. Such criteria ended up yielding 181 well-matched patient pairs for comparative analysis of procedural, echocardiographic, and clinical outcomes for patients.

Prior to score matching there were several notable differences between the various treatment groups and patient populations. Patients who received the S3UR valve were slightly younger on average (71.00 ± NA vs. 81.60 ± 9.87 years). Sex distribution was similar (S3UR: 64% male vs. S3U/S3: 63%), however, there was to note a slightly lower tobacco use rate in the S3UR group (45% vs. 50%; ASD = 0.095). Additionally, preoperative echocardiographic parameters as we know it such as left ventricular ejection fraction (PreOP_LVEF) were relatively similar S3UR: 57.97 ± 12.37% vs. S3U/S3: 59.62 +/-9.98%, and preoperative mean gradient (PreOP_MG) was modestly lower in the S3UR group (35.54 ± 12.64 mmHg vs. 36.65 ± 13.37 mmHg; ASD = 0.085). Other differences were observed in rates of preexisting atrial fibrillation (31% vs. 30%; ASD = 0.025) and presence of moderate/severe tricuspid regurgitation (47% vs. 46%; ASD = 0.014). There was also a difference observed in functional status as measured by the Kansas City Cardiomyopathy Questionnaire (KCCQ) (KCCQ12_Overall: 56.77 ± 29.38 in S3UR vs. 54.46 ± 28.08 in S3U/S3).

Following matching, however, the cohorts were known to achieve excellent balance across all key variables and data. The matched mean age was 81.3 ± 9.6 years in the S3UR group and 81.0 ± 9.9 years in the S3U/S3 group, with an absolute standardized difference (ASD) of 0.034; the distribution of sex also remained consistent (S3UR: 63% male vs. S3U/S3: 61%; ASD = 0.034). Additionally, STS risk scores were similar (5.18 ± 4.54 for S3UR vs. 4.72 ± 4.10 for S3U/S3; ASD = 0.108), as were left ventricular ejection fraction values (58.97 ± 11.03% vs. 58.14 ± 9.30%; ASD = 0.081). Key echocardiographic parameters such as preoperative aortic valve area (0.80 ± 0.17 cm^2^ vs. 0.78 ± 0.18 cm^2^; ASD = 0.12) and AV peak gradient (60.76 ± 20.19 mmHg vs. 58.33 ± 20.34 mmHg; ASD = 0.026) were also well matched and similar across patient groups. It is to note minor imbalances persisted in variables such as the number of prior cardiac surgeries (0.13 ± 0.37 vs. 0.17 ± 0.38; ASD = 0.103), presence of postprocedural aortic regurgitation (PP_AR: 5% vs. 3%; ASD = 0.115), and diabetes mellitus (44% vs. 41%; ASD = 0.056), however, these remained within an acceptable range and were therefore not significant.

Importantly, all matched variables had ASDs less than 0.15, and the majority were well below the 0.1 threshold, which indicates a strong covariate balance and also minimizes the risk of residual confounding as would be a negative consequence. This methodological rigor also supports the internal validity of outcome comparisons between the S3UR and S3U/S3 groups in subsequent analyses and studies outside of UCLA.

### Procedural Outcomes

Procedural outcomes for the propensity score–matched cohort are summarized in Table 2. Transfemoral access was the dominant approach in both groups (S3UR: 92.9%, S3U/S3: 93.7%) and no conversions to alternative access or surgical intervention were required. The majority of procedures were elective (S3UR: 87.0%, S3U/S3: 89.9%) but there were to note slightly more urgent procedures performed in the S3UR group (13.0% vs. 10.1%).

**Table 1.** Baseline Characteristics for the Unmatched and Matched Cohorts.

|  | Unmatched Cohort |  |  | Matched Cohort |  |  |
| --- | --- | --- | --- | --- | --- | --- |
|  | Unmatched - S3/S3U | Unmatched - S3UR | Unmatched - ASD | Matched - S3/S3U | Matched - S3UR | Matched - ASD |
| Age | 81.60 ± 9.87 | 71.00 ± nan | | 81.32 ± 9.61 | 71.00 ± nan | |
| Sex | 187 (63%) | 124 (64%) | 0.013 | 111 (61%) | 114 (63%) | 0.034 |
| STSRiskScoreValue | 4.49 ± 4.01 | 5.36 ± 4.66 | 0.2 | 4.72 ± 4.10 | 5.18 ± 4.54 | 0.108 |
| TobaccoUse | 140 (50%) | 82 (45%) | 0.095 | 80 (48%) | 77 (45%) | 0.058 |
| PreOP_LVEF | 59.62 ± 9.98% | 57.98 ± 12.37 | 1.1 | 58.14 ± 9.30 | 58.97 ± 11.03 | 0.081 |
| PreOP_MG | 36.65 ± 13.37 | 35.54 ± 12.64 | 0.085 | 34.97 ± 12.91 | 36.20 ± 12.52 | 0.097 |
| PP_AR | 12 (4%) | 10 (5%) | 0.052 | 5 (3%) | 9 (5%) | 0.115 |
| PreOP_AVA | 0.77 ± 0.21 | 0.80 ± 0.18 | 0.18 | 0.78 ± 0.18 | 0.80 ± 0.17 | 0.12 |
| NumPrevCardSurg | 0.16 ± 0.41 | 0.14 ± 0.37 | 0.052 | 0.17 ± 0.38 | 0.13 ± 0.37 | 0.103 |
| HmO2 | 29 (10%) | 16 (8%) | 0.056 | 14 (8%) | 15 (8%) | 0.02 |
| KCCQ12_Overall | 54.46 ± 28.08 | 56.77 ± 29.38 | 0.08 | 54.79 ± 23.89 | 56.27 ± 29.00 | 0.056 |
| Atrial Fibrillation | 88 (30%) | 60 (31%) | 0.025 | 34 (19%) | 56 (31%) | 0.281 |
| Carotid Artery Stenosis | 14 (5%) | 8 (4%) | 0.03 | 9 (5%) | 8 (4%) | 0.026 |
| Cerebrovascular Disease | 50 (17%) | 5 (3%) | 0.482 | 7 (4%) | 5 (3%) | 0.062 |
| Chronic Lung Disease | 0.32 ± 0.47 | 0.23 ± 0.42 | 0.211 | 0.15 ± 0.38 | 0.24 ± 0.43 | 0.206 |
| Diabetes Mellitus | 146 (49%) | 87 (45%) | 0.091 | 79 (44%) | 74 (41%) | 0.056 |
| FiveMWTTime | 6.94 ± 2.25 | 7.60 ± 3.32 | 0.234 | 7.38 ± 1.93 | 7.21 ± 2.52 | 0.076 |
| BMI | 27.37 ± 5.99 | 26.77 ± 5.03 | 0.109 | 27.57 ± 5.94 | 26.87 ± 5.10 | 0.127 |
| **Echo data** |  |  |  |  |  |  |
| PreprocMR | 136 (46%) | 109 (56%) | 0.202 | 107 (59%) | 99 (55%) | 0.089 |
| PreprocTR | 138 (46%) | 92 (47%) | 0.014 | 76 (42%) | 83 (46%) | 0.078 |
| AVPeakGrad | 61.46 ± 21.23 | 59.73 ± 20.36 | 0.083 | 58.33 ± 20.34 | 60.76 ± 20.19 | 0.12 |
| ProOP_AVA | 0.77 ± 0.21 | 0.80 ± 0.18 | 0.18 | 0.78 ± 0.18 | 0.80 ± 0.17 | 0.12 |

**Table 2:** Procedure Outcomes for the Matched Cohort.

|  | S3UR Fraction | S3UR Mean (%) | S3UR SD (%) | S3/S3U Fraction | S3/S3U Mean (%) | S3/S3U SD (%) |
| --- | --- | --- | --- | --- | --- | --- |
| Transfemoral Access | 143/154 | 92.9 | 25.8 | 74/79 | 93.7 | 24.5 |
| Elective Procedure | 134/154 | 87 | 33.7 | 71/79 | 89.9 | 30.4 |
| Urgent Procedure | 20/154 | 13 | 33.7 | 8/79 | 10.1 | 30.4 |
| General Anesthesia | 30/154 | 19.5 | 39.7 | 11/79 | 13.9 | 34.8 |
| Conscious Sedation | 124/154 | 80.5 | 39.7 | 68/79 | 86.1 | 34.8 |
| Procedure aborted | 0/154 | 0 | 0 | 0/79 | 0 | 0 |
| Annular rupture | 0/154 | 0 | 0 | 0/79 | 0 | 0 |
| Aortic dissection | 0/154 | 0 | 0 | 0/79 | 0 | 0 |
| Coronary obstruction | 0/154 | 0 | 0 | 0/79 | 0 | 0 |
| Device embolization | 0/154 | 0 | 0 | 0/79 | 0 | 0 |
| Perforation + tamponade | 0/154 | 0 | 0 | 0/79 | 0 | 0 |
| Implantation success | 18/154 | 11.7 | 32.2 | 15/79 | 19 | 39.5 |
| Multiple complications | 18/154 | 11.7 | 32.2 | 15/79 | 19 | 39.5 |
| Discharged Home Alive | 153/154 | 99.4 | 8.1 | 78/79 | 98.7 | 11.3 |
| Valve size: | S3UR (%) | S3/S3U (%) |  |  |  |  |
| 26 mm | 41.6 | 39.7 |  |  |  |  |
| 23 mm | 29.9 | 35.9 |  |  |  |  |
| 29 mm | 27.9 | 24.4 |  |  |  |  |
| 20 mm | 0.6 | 0 |  |  |  |  |

General anesthesia was notably used in 19.5% of S3UR cases compared to 13.9% in the S3U/S3 group, while conscious sedation was used more frequently in S3U/S3 patients (86.1% vs. 80.5%), which could be linked to a change in overall hospital protocol. Additionally, perhaps due to the single-center nature of this study, no cases of procedural abortion, annular rupture, aortic dissection, coronary obstruction, device embolization, or cardiac perforation were observed in either cohort.

Valve size selection was similar between groups. The 26 mm valve was most commonly implanted (S3UR: 41.6%, S3U/S3: 39.7%), followed by 23 mm (S3UR: 29.9%, S3U/S3: 35.9%) and 29 mm (S3UR: 27.9%, S3U/S3: 24.4%). The 20 mm valve was used only once in the S3UR group (0.6%) and not at all in the S3U/S3 group. Implantation success was also high in both groups (S3UR: 98.7%, S3U/S3: 98.1%), and 99.4% of S3UR patients and 98.7% of S3U/S3 patients were discharged home without major complications from their respective procedures or devices.

### In-Hospital and 30-Day Clinical Outcomes

In-hospital and 30-day clinical outcomes for the matched cohort are presented in Table 3. Event rates during the index hospitalization were quite low in both groups and we note there were no statistically significant differences in all-cause mortality (S3UR: 0.65% vs S3U/S3: 1.27%; p=0.663), cardiac death (0.65% vs 0%; p=0.316), or stroke (0.65% vs 1.27%; p=0.663). Additionally, the requirement for new permanent pacemaker implantation was slightly higher in the S3UR group (1.3%) compared to S3U/S3 (1.27%) though not statistically significant (p=0.983). New-onset atrial fibrillation, aortic valve reintervention, life-threatening bleeding, and major vascular complications were not observed in either group during the index hospitalization or 30-day follow-up.

**Table 3:** In-Hospital and 30-Day Clinical Outcomes for Matched Cohort.

| Table 3: In-Hospital and 30-Day Clinical Outcomes for Matched Cohort |  |  |  |  |  |  |
| --- | --- | --- | --- | --- | --- | --- |
|  | In-hospital |  |  | 30-Day |  |  |
| Outcome | S3/S3U | S3UR | Z-Test P Value | S3/S3U % ± SD | S3/S3U n/N | P-Value (Z-Test) |
| All-cause death | 1.3 ± 1.26% (1/79) | 0.6 ± 0.65% (1/154) | 0.629 | 1.27 ± 1.26 | 1/79 | 0.663 |
| Cardiac death | 1.3 ± 1.26% (1/79) | 0.6 ± 0.65% (1/154) | 0.629 | 0.00 ± 0.00 | 0/79 | 0.3157 |
| Stroke | 1.3 ± 1.26% (1/79) | 1.3 ± 0.91% (2/154) | 0.983 | 1.27 ± 1.26 | 1/79 | 0.663 |
| New pacemaker | 2.5 ± 1.77% (2/79) | 5.2 ± 1.79% (8/154) | 0.342 | 1.27 ± 1.26 | 1/79 | 0.9831 |
| New-onset atrial fibrilla | 5.1 ± 2.47% (4/79) | 2.6 ± 1.28% (4/154) | 0.328 | 0.00 ± 0.00 | 0/79 | 0 |
| Aortic valve reinterven | 0.0 ± 0.00% (0/79) | 0.0 ± 0.00% (0/154) | 1 | 0.00 ± 0.00 | 0/79 | 0 |
| Readmission | | | | 13.92 ± 3.90 | 11/79 | 0.1686 |
| Life-threatening bleed | 2.5 ± 1.77% (2/79) | 4.5 ± 1.68% (7/154) | 0.45 | 0.00 ± 0.00 | 0/79 | 0 |
| Major vascular complic | 1.3 ± 1.26% (1/79) | 1.3 ± 0.91% (2/154) | 0.983 | 0.00 ± 0.00 | 0/79 | 0 |

Readmission within 30 days occurred in 7.79% of patients in the S3UR group compared to 13.92% in the S3U/S3 group, though this difference was not statistically significant (p=0.169).

### Echocardiographic Outcomes

| Label | Mean (S3/S3U) | SD (S3/S3U) | Fraction (S3/S3U) | Mean (S3UR) | SD (S3UR) | Fraction (S3UR) | P-Value (S3UR) |
| --- | --- | --- | --- | --- | --- | --- | --- |
| EOA, cm<sup>2</sup> (1-Year) | 1.138888889 | 0.3507361872 | N/A | 1 | N/A | N/A | N/A |
| EOA, cm<sup>2</sup> (30-Day) | 1.112676056 | 0.4644288589 | N/A | 1.308510638 | 0.5292842752 | N/A | 0.01158984912 |
| EOA, cm<sup>2</sup> (Discharge) | 1.152777778 | 0.5479725043 | N/A | 1.454545455 | 0.6135474399 | N/A | 0.000253512861 |
| Mean gradient ≥20 mm Hg, % (1-Year) | 10.10% | 30.20% | 8/79 | 0.00% | 0.00% | 0/154 | 5.85E-05 |
| Mean gradient ≥20 mm Hg, % (30-Day) | 8.90% | 28.40% | 7/79 | 0.00% | 0.00% | 0/154 | 0.000176297566 |
| Mean gradient ≥20 mm Hg, % (Discharge) | 2.50% | 15.70% | 2/79 | 1.30% | 11.30% | 2/154 | 0.4927893433 |
| Mean gradient, mm Hg (1-Year) | 14.63888889 | 6.870860947 | N/A | 6 | N/A | N/A | N/A |
| Mean gradient, mm Hg (30-Day) | 12.52702703 | 5.187505994 | N/A | 9.057692308 | 2.935642776 | N/A | 2.08E-07 |
| Mean gradient, mm Hg (Discharge) | 10.21794872 | 5.052059978 | N/A | 7.452054795 | 3.37349137 | N/A | 1.39E-05 |
| Paravalvular leak - Mild (1-Year) | 58.80% | 49.20% | 10/17 | 0.00% | 0.00% | 0/0 | 1 |
| Paravalvular leak - Mild (30-Day) | 100.00% | 0.00% | 6/6 | 100.00% | 0.00% | 12/12 |  |
| Paravalvular leak - Mild (Discharge) | 25.00% | 43.30% | 2/8 | 25.00% | 43.30% | 3/12 | 1 |
| Paravalvular leak - Moderate (1-Year) | 0.00% | 0.00% | 0/17 | 0.00% | 0.00% | 0/0 | N/A |
| Paravalvular leak - Moderate (30-Day) | 0.00% | 0.00% | 0/6 | 0.00% | 0.00% | 0/12 | N/A |
| Paravalvular leak - Moderate (Discharge) | 12.50% | 33.10% | 1/8 | 8.30% | 27.60% | 1/12 | 0.7609067271 |
| Paravalvular leak - None (1-Year) | 0.00% | 0.00% | 0/17 | 0.00% | 0.00% | 0/0 | N/A |
| Paravalvular leak - None (30-Day) | 0.00% | 0.00% | 0/6 | 0.00% | 0.00% | 0/12 | N/A |
| Paravalvular leak - None (Discharge) | 0.00% | 0.00% | 0/8 | 0.00% | 0.00% | 0/12 | N/A |
| Paravalvular leak - Severe (1-Year) | 0.00% | 0.00% | 0/17 | 0.00% | 0.00% | 0/0 | N/A |
| Paravalvular leak - Severe (30-Day) | 0.00% | 0.00% | 0/6 | 0.00% | 0.00% | 0/12 | N/A |
| Paravalvular leak - Severe (Discharge) | 0.00% | 0.00% | 0/8 | 0.00% | 0.00% | 0/12 | N/A |

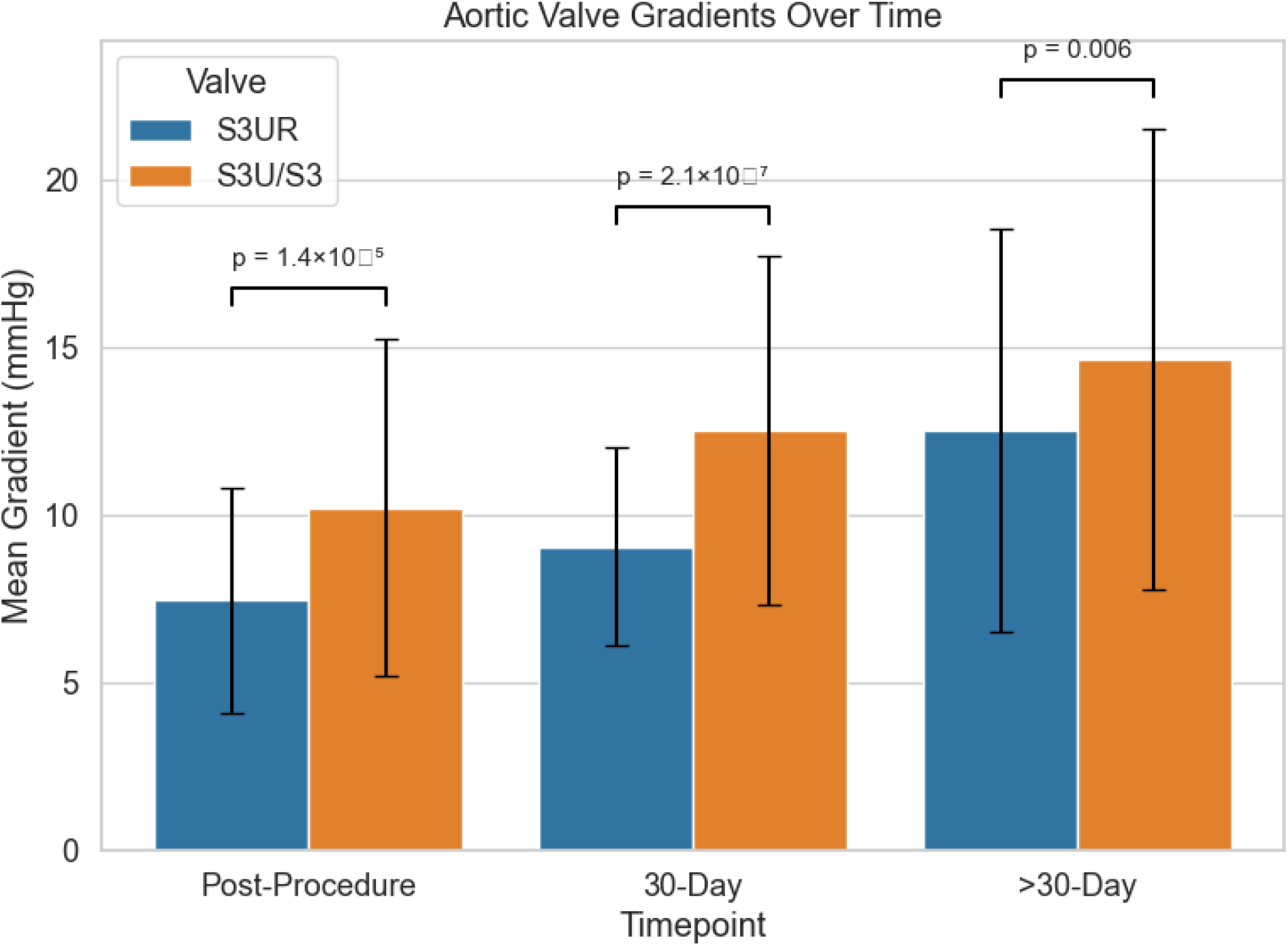

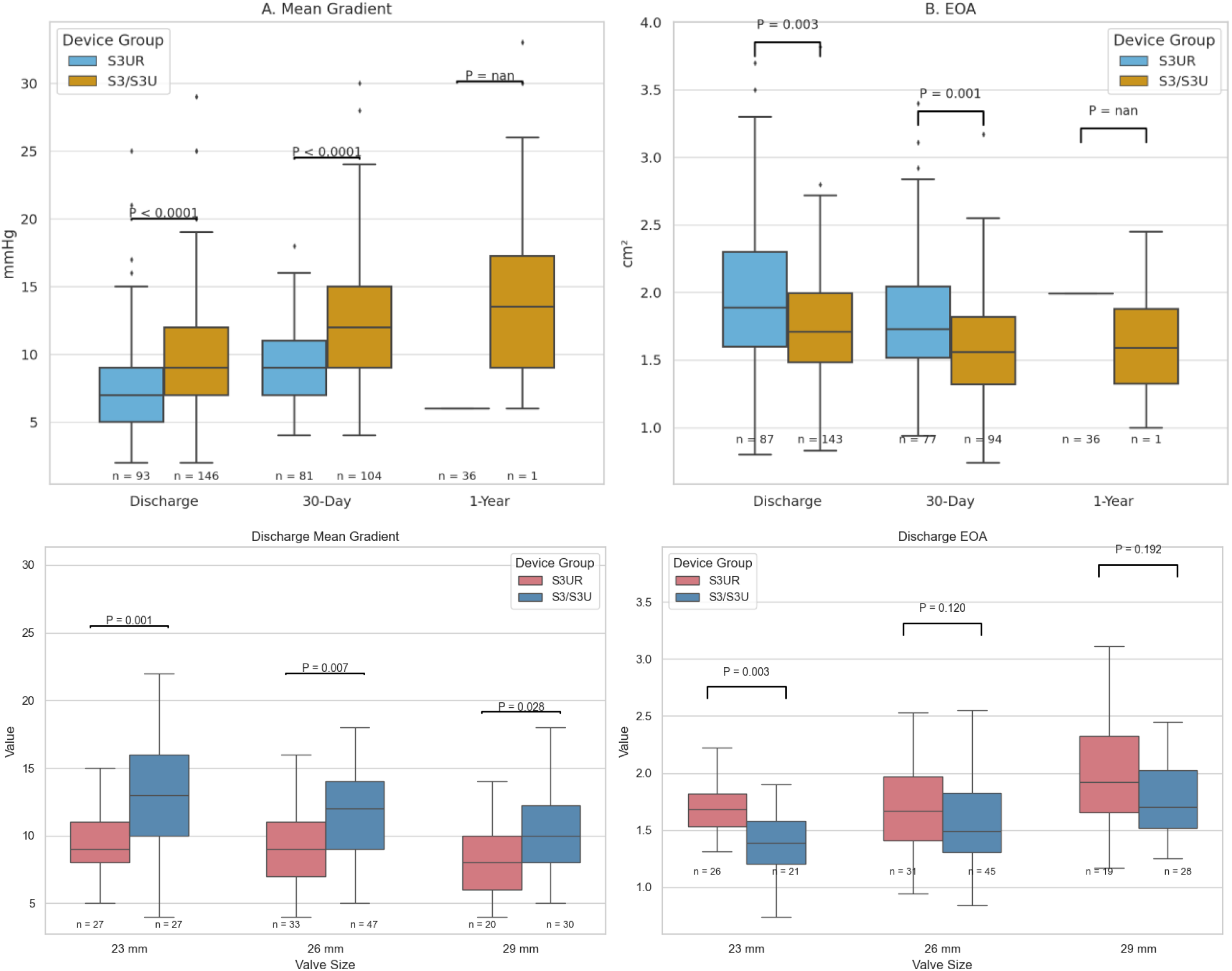

Echocardiographic results for the matched UCLA cohort are summarized in Table 4. Post-procedure (PP) mean aortic valve gradients were significantly lower in patients receiving the S3UR valve compared to those treated with the S3U/S3 platform (S3UR: 7.45 ± 3.37 mmHg vs. S3U/S3: 10.22 ± 5.05 mmHg; p = 1.39 × 10^−5^), indicating better immediate hemodynamic performance.

At 30-day follow-up, this trend persisted, with the S3UR group maintaining significantly lower mean gradients (S3UR: 9.06 ± 2.94 mmHg vs. S3U/S3: 12.53 ± 5.19 mmHg; p = 2.08 × 10−^7^). Beyond 30 days (F2_F) we note that mean gradients remained lower in the S3UR cohort (12.53 ± 6.00 mmHg vs. 14.64 ± 6.87 mmHg; p = 0.006), which supported sustained valve performance. Paravalvular leak (PVL) rates were notably lower with the S3UR valve and no patients in the S3UR group experienced moderate or greater aortic regurgitation at any time point; however, moderate PVL was observed in the S3U/S3 cohort at both the 30-day mark (8.9%) and longer-term follow-up (10.1%) (p = 5.85 × 10^−5^ for both). Post-procedural moderate AR was also lower with S3UR (1.3% vs. 2.5%, p = 0.49), although this did not reach the point of statistical significance and therefore could be negligible.

To analyze, this data demonstrate that S3UR provided superior post-procedural and follow-up valve hemodynamics with a markedly lower incidence of paravalvular regurgitation when compared to S3U/S3 at Ronald Raegan.

## Discussion

This study represents one of the first institution-specific evaluations of the fifth-generation balloon-expandable Sapien 3 Ultra Resilia (S3UR) valve in a matched TAVR population and thus provides important novel insights into its procedural, clinical, and hemodynamic performance in a real-world setting. Our findings build upon national data from the STS/ACC TVT Registry and thus confirm that the S3UR valve offers incremental benefits in terms of valve performance while maintaining procedural safety and also clinical efficacy in this case.

The most salient finding of this analysis was the superior hemodynamic profile of the S3UR valve, which was effectively evidenced by significantly lower mean aortic gradients both immediately post-procedure and at 30-day and >30-day follow-up. This improvement as depicted likely reflects a combination of factors which include the revised leaflet suspension mechanism in smaller valve sizes, the Resilia anti-calcification tissue treatment, and the novel enhanced skirt design that also improves coaptation and reduces residual PVL within newer patients.

These notable structural modifications have translated into tangible clinical benefits: in our cohort, no patients with S3UR experienced moderate or greater paravalvular regurgitation at any follow-up timepoint, compared to 8.9– 10.1% in the matched S3U/S3 group. This observation is consistent with the TVT registry analysis where S3UR also demonstrated a lower incidence of moderate or greater PVL (0.6% vs. 1.3%, p < 0.01).

While most procedural metrics were similar between groups, it is to note that S3UR cases showed slightly higher rates of general anesthesia as well as urgent procedures. These differences are likely a reflection of random patient selection and institutional workflow rather than actual device characteristics and procedural outcomes. Importantly, procedural safety was excellent across both groups with no cases of annular rupture, device embolization, or conversion to open surgery within the matched cohort. Implantation success also exceeded 98% in both groups with nearly all patients being discharged home safely, reinforcing the platform’s adaptability to enhanced recovery and minimalist TAVR protocols.

The S3UR group also had a numerically higher, though importantly statistically nonsignificant, rate of new permanent pacemaker implantation (5.2% vs. 2.5%). While this trend mirrors observations in some multicenter reports done, it contrasts with early studies suggesting comparable conduction profiles between S3UR and its predecessor [1] Given that pacemaker risk is multifactorial meaning it is often driven by implantation depth, membranous septum length, and baseline conduction disease, we must have future investigations clarify whether valve design accounts for this variation or if the changes are more likely due to operator technique within the procedure.

Readmission rates at 30 days were lower in the S3UR group (7.8% vs. 13.9%), though this did not reach statistical significance. This may reflect improved post-discharge hemodynamics and fewer PVR-related symptoms, though sample size limits definitive interpretation.

In terms of limitations, this is a single-center retrospective analysis, subject to unmeasured confounding despite careful matching. Echocardiographic measurements were site-reported and also therefore may be subject to interobserver variability. Finally, long-term durability and structural valve deterioration were not evaluated due to the fact that many of the newer patients in 2024 have not been reevaluated for their post one year follow up and ongoing surveillance will be necessary to confirm the durability benefits of Resilia tissue in such populations.

Despite these limitations, our study adds to the growing body of evidence supporting the use of the S3UR platform and demonstrates its consistent performance in a high-volume academic center across a wide range of patient anatomies and clinical presentations.

## Conclusion

In this matched, single-center cohort, the Sapien 3 Ultra Resilia valve demonstrated excellent procedural success, low complication rates, and significantly improved hemodynamic performance compared to the previous-generation

S3U/S3 valves. Perhaps the most striking clinical advantage as depicted in this study was the complete absence of moderate or greater paravalvular regurgitation in S3UR recipients at all timepoints, reflecting the effectiveness of the updated skirt design and valve-tissue interface improvements.

While mortality, stroke, and major vascular complications remained low and comparable between groups, the S3UR valve showed early evidence of more favorable readmission rates and sustained lower aortic gradients, suggesting potential long-term clinical and quality-of-life benefits.

These findings support the routine use of the S3UR valve in contemporary TAVR practice and reinforce its role as a default platform in transfemoral procedures at experienced centers. Further longitudinal data will be essential to assess long-term durability and to clarify whether the early hemodynamic advantages observed with S3UR translate into improved structural valve longevity and clinical outcomes over time.

## Data Availability

The data supporting the findings of this study are available from the corresponding author upon reasonable request. Due to institutional review board restrictions and the inclusion of protected health information, the data are not publicly available.

## Notes

### Competing Interest Statement

The authors have declared no competing interest.

### Funding Statement

None was recieved.

### Author Declarations

UCLA med: IRB-16-0680-AM-021

